# Family supplemented patient monitoring after surgery (SMARTER): process evaluation of a complex intervention in Uganda

**DOI:** 10.1101/2024.05.24.24307741

**Authors:** Adam Hewitt-Smith, Fred Bulamba, Juliana Nanimambi, Lucy Ruth Adong, Bernard Emacu, Mary Kabaleta, Justine Khanyalano, Maiga Ayub Hussein, Charles Mugume, Joanitah Nakibuule, Loretta Nandyose, Martin Sejja, Winfred Weere, Rupert M. Pearse, Timothy Stephens

## Abstract

**Background:** Improving outcomes after surgery in Africa is a priority. The SMARTER Pilot Trial was a step-wedge cluster-randomised trial of family member supplemented vital signs monitoring after surgery. A concurrent process evaluation provides contextual understanding of intervention delivery.

**Methods:** Mixed methods approach with qualitative data sources including field notes from a research team diary and focus group discussions. Deductive analysis used the consolidated framework for implementation research. Quantitative data evaluating the efficacy of family members recognising abnormal vital signs and reporting them to nursing staff were collected following a prespecified intervention review held after two months.

**Findings:** Focus group discussions were conducted with 16 nurses and research assistants. Field notes included 88 episodes documented throughout the trial in a research team diary. Quantitative data were collected in the final 397 patients following ethics amendments. Intervention facilitators included: relative advantage, inner context factors including tension for change and relative priority, and individual characteristics centred around knowledge and beliefs. Available resources, culture, and compatibility were identified as important barriers, with a smaller negative influence from self-efficacy and intervention complexity. Family members recognised 91.3% (42/46) of abnormal sets of vital signs and communicated 100% (42/42) of these to a member of the nursing or medical team. The team responded 90.5% (38/42) of the time.

**Interpretation:** Family members were able to supplement nurse led monitoring of patients after surgery. This complex intervention was affected by context specific positive and negative influences. Scaling this intervention requires careful consideration of local context during planning.

**Trial registration:** SMARTER Pilot Trial registered on clinicaltrials.gov - NCT04341558

## Introduction

Compared to global averages, the typical surgical patient in Africa is twice as likely to die, despite being younger and with fewer comorbidities.^1^ The majority of these deaths occur on the hospital wards after surgery.^1-3^ These deaths represent ‘failure to rescue’ i.e. death of a patient following unrecognised physiological deterioration, caused by a perioperative complication.^4,5^ Failure to appropriately monitor patients, failure to recognise their physiological deterioration, and failure to act when deterioration occurs are significant contributors to rates of failure to rescue.^6-9^ Across Africa, with patient to nurse ratios of up to 60:1, workforce shortages make close postoperative monitoring difficult to achieve.^10^ The SMARTER cluster-randomised pilot trial tested the efficacy of an intervention to train family members to perform basic vital signs monitoring, supplementing routine postoperative monitoring by staff in a regional hospital in Uganda. [submitted for publication]

Healthcare delivery, with its multiple interacting components is naturally complex. Whilst all healthcare environments are complex, the variability in healthcare delivery in low-income settings makes them much more unpredictable.^11,12^ This contextual understanding is important when developing or testing interventions to allow effective translation of evidence into day-to-day clinical practice. Pragmatic interventions are also often complex themselves. Training family members to perform vital signs monitoring on inpatient wards in hospital relies on multiple interacting factors to be effective. This is in direct comparison to the meticulous environment that limits confounders in a randomised controlled trial for example, when testing the effectiveness of a novel drug. Process evaluations are studies that run alongside an intervention trial to help understand the context underlying the intervention’s impact. Frameworks and guidance exist to enable researchers to co-design process evaluations in accordance with best practice.^13,14^ Following this guidance, we conducted a process evaluation of the SMARTER Pilot Trial.

In this paper, we describe our mixed methods process evaluation. This allows a greater contextual interpretation of the pilot trial results. It enhances our understanding of the complex healthcare environment in eastern Uganda and provides important feedback that can be incorporated into the design of future trials or efforts to scale interventions that involve family members in healthcare settings.

## Methods

### SMARTER pilot trial

The SMARTER Pilot trial was a stepped-wedge cluster-randomised pilot trial evaluating a complex intervention training family members to support nursing staff to take and record patients’ vital signs after surgery. This intervention was developed from programme theory where increased postoperative monitoring leads to earlier recognition of deterioration, an earlier response and therefore reduction in the number of postoperative deaths. (Figure 1) The primary outcome was the frequency of vital signs from arrival on the postoperative ward to the end of the third postoperative day. Full details of the trial and results are reported elsewhere. [submitted for publication]

**Figure 1.**
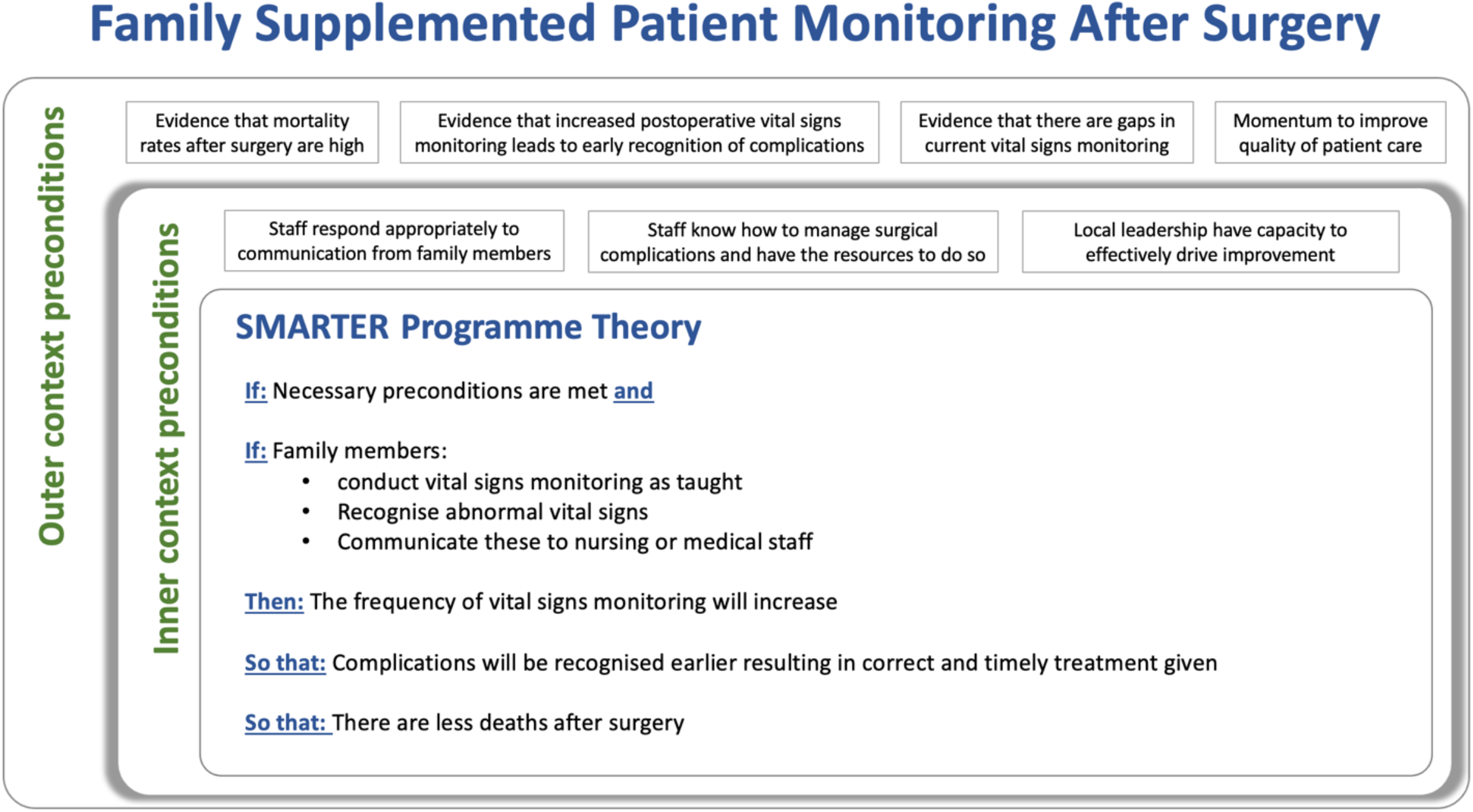
SMARTER programme theory

### Study design

We undertook a prospectively planned mixed methods process evaluation. The pilot trial design included a predetermined point at two months for a formal evaluation of the intervention to enable further intervention refinement and adaption. Focus group discussions with ward nurses and research assistants were conducted at two months and at trial completion. Field notes were documented throughout the trial by the research team in a diary. Following the prespecified review at two months and ethics approval of the amended protocol, additional quantitative data was collected on all remaining patients in the trial to evaluate the effectiveness of the intervention. On a daily basis the research team used the family member observation chart to identify abnormal vital signs from the previous 24-hour period. Variables included the number of sets of abnormal vital signs, whether these were recognised by the family member and communicated to a member of staff, and whether the staff responded. All data collection was completed without knowledge of the trial results. Ethics approval was granted by Mbale Regional Referral Hospital (RRH) Research Ethics Committee (MRRH-2020-7), Uganda National Council for Science & Technology (HS944ES) and the Queen Mary Ethics of Research Committee (QMERC2019/72).

### Data analysis

Data from the field notes and focus group discussions were manually and independently analysed by TS and AHS using a deductive approach.^15^ Thematic coding was based upon the constructs defined in the Consolidated Framework for Implementation Research (CFIR).^16^ The CFIR provides a mutual language that can be used across different complex environments to help organise and understand common constructs, focusing on five areas: the inner and outer context, the implementation, individuals involved and the intervention itself. Data sources were initially read from start to finish to familiarise ourselves with the content as a whole. Data was then re-read to identify fragments of text (sentences or small groups of sentences) that corresponded to identified constructs from within the CFIR. Regular meetings were held to discuss and agree on the coding decisions. Quantitative data was analysed using Microsoft Excel (Microsoft Corporation, 2018) and is presented using descriptive statistics.

## Results

### Main trial findings

The SMARTER pilot trial included 1395 patients across four clusters over a six-month period between April to October 2021. There were 12.5 times as many sets of vital signs in the intervention group when compared to the usual care group. (Incident rate ratio, intervention vs usual care, 12.5 [8.9 - 17.7], p=0.001). [ref]

### Intervention efficacy

Quantitative process evaluation data was collected for a total of 397 patients over 783 days. 46/783 (5.9%) abnormal sets of vital signs were documented during this time. Of these, 91.3% (42/46) were recognised by family members, 100% (42/42) were communicated to a member of the nursing or medical team and the team responded 90.5% (38/42) of the time (Figure 1).

### Factors influencing intervention delivery

Focus group discussions with 16 ward nurses and research assistants were conducted at two months and at trial completion. Field notes included 88 episodes documented throughout the trial in a research team diary and in a prespecified free text box on the case report form. Delivery of the SMARTER intervention during the pilot trial was influenced by factors that can be categorised into two major groups, facilitators (those with a positive influence) and barriers (those with a negative influence). Table 1 defines the constructs from the CFIR framework that these influences fall under and illustrates corresponding quotations for each.

**Table 1.** Barriers and facilitators identified during the process evaluation with their indicative quotations.

| Domain | Construct | Short description of construct | Indicative quotations |
| --- | --- | --- | --- |
| ***Facilitators (positive influences)*** |  |  |  |
| Intervention characteristics | Relative advantage | Stakeholders' perception of the advantage of implementing the intervention versus an alternative solution | *"I feel the programme has relieved us of some work, if the attendant is in position to come and tell you something has gone wrong, that means they have really helped you, than coming back in the morning after some rest and find when the patient is either deteriorated, you have to resuscitate or the patient has passed on."* [Nurse, FGD 2] |
| Inner setting | Tension for change | The degree to which stakeholders perceive the current situation as intolerable or needing change. | *"When a patient has just come from theatre, we need to first take vitals, they have requested half hourly.....when you are alone on the ward with many patients, you will hardly meet that because you can't just be on one patient."* [Nurse, FGD 2] |
|  | Relative priority | Individuals' shared perception of the importance of the implementation within the organization | *A family member who had finished day 3 [of the] intervention the previous day requested to continue close monitoring on his patient since the results were so useful in the management of the patient.*<br>[Field notes] |
| Characteristics of individuals | Knowledge and beliefs about the intervention | Individuals' attitudes toward and value placed on the intervention as well as familiarity with facts, truths, and principles related to the intervention. | *".....there is some knowledge being spread in the community. I am sure those people who have learnt to do something on the ward, they are already telling others, when you go to hospital you do not only go to look after your patient but there is also some knowledge imparted in you by the nurses."* [Nurse, FGD 2] |
|  | Other personal attributes | A broad construct to include other personal traits such as tolerance of ambiguity, intellectual ability, motivation, values, competence, capacity, and learning style. | *one patient had an illiterate attendant stay overnight. However, a neighbouring attendant had just finished [the] intervention for his patient, so he was willing to help the attendant to do vitals for his patient.* [Field notes] |

| **Barriers (negative influences)** |  |  |  |
| --- | --- | --- | --- |
| Inner setting | Available resources | The level of resources dedicated for implementation and on-going operations, including money, training, education, physical space, and time. | *While taking vital signs, a caretaker recorded oxygen levels less than 40% so she tried to look for a nurse on duty but couldn't see her. She had to just look on to see what happened to her patient. [Field notes]* |
|  | Culture | Norms, values, and basic assumptions of a given organization. | *The carers who identify abnormal vital signs at night do not alert the medical team because according to them 'we fear waking up the nurses at night to destroy their sleep'. [Field notes]* |
|  | Compatibility | The degree of tangible fit between meaning and values attached to the intervention by involved individuals, how those align with individuals' own norms, values, and perceived risks and needs, and how the intervention fits with existing workflows and systems. | *Attendants were asked to stay outside. They were unable to do vitals, and this also affected our training for new participants. [Field notes]* |
| Intervention characteristics | Complexity | Perceived difficulty of implementation, reflected by duration, scope, radicalness, disruptiveness, centrality, and intricacy and number of steps required to implement | *[I] noted that our vital signs chart is giving some headache to our participants. Why? Because on the monitors, oxygen level comes first then heart rate yet on our vital signs sheet heart rate comes first the oxygen last. Therefore, you find that some patients may end up recording the oxygen level figures on the heart rate. [Field notes]* |
| Characteristics of individuals | Self efficacy | Individual belief in their own capabilities to execute courses of action to achieve implementation goals. | *"...they fear they are going to do work they don't understand, some of them have never been to school so what they do is just ignore the pen... ...i think the issue is about reading and understanding" [RA, FGD 1]* |

### Facilitators

Relative advantage [Characteristics of the intervention] was the construct most commonly coded across all data sources. Healthcare workers, research staff, patients and their families shared the perception that the SMARTER intervention offered a significant advantage over the alternative of changing nothing. Nursing staff on the wards saw first-hand how the intervention helped to relieve them of some of their workload and at the same time contributed to improving the quality of care. This perception had a strong positive influence on how the nursing staff engaged with the intervention. They trusted the ability of the family members to monitor the vital signs and alert them when there was a problem.

*“It actually improved, because now say for instance you are one nurse on the ward, when the caretaker is able to record the vitals and she knows when she or he [has to] come to the nurse, it gives me time to attend to other patients knowing that by the time the other attendant comes that means there is a problem there. And if I am to check you find they have already recorded*.*”* [Nurse, FGD 2]

Tension for change [inner setting] is a driving force that often leads to changes in practice. Healthcare settings in low-income countries commonly face significant human resource shortages. Nursing staff in hospital wards can be overwhelmed with competing pressures that force them to prioritise what they spend their time doing. This tension for change was evident during the SMARTER Pilot Trial where a single nurse on the postnatal ward had responsibility for helping with the ward round, delivering prescribed treatment, writing discharges for patients after vaginal deliveries, teaching mothers about post discharge care, collecting new admissions from labour suite theatre and monitoring patients after surgery. Staff on the wards know they need to monitor their patients closely after surgery but when they are alone there is simply not the time.

Relative priority [inner setting] and individuals’ knowledge and beliefs [characteristics of individuals] were the next most commonly coded constructs displaying a positive influence on the intervention. Patients and their family members in particular, recognised the importance of close monitoring. At times during the trial, patients who did not meet the inclusion criteria also wanted to be closely monitored and made special requests to be trained.

*One of the patients who was operated on was not recruited…today one of his attendants approached us and said he should be trained, consented, and included in the study. I explained to him why he could not be included in the study, but he still insisted. [Field notes]*

The impact of an intervention that involves members of the family or community is often further reaching than initially expected. Increased positive community awareness of work in the hospital encourages patients to attend earlier and improves community engagement in their care.

*“……there is some knowledge being spread in the community. I am sure those people who have learnt to do something on the ward, they are already telling others, when you go to hospital you do not only go to look after your patient but there is also some knowledge imparted in you by the nurses*.*”* [Nurse, FGD 2]

Variation in behaviours and attitudes of the general public in a hospital is expected. Patients and their families come from many different backgrounds. Even when some family members were hesitant to take on the extra responsibility of monitoring their patients closely, the patients themselves were enthusiastic. This was also seen when their family members were illiterate and unable to participate.

*“the perception of the patients is very good, very positive because this is a person who is very eager to know his physiology is running. The attendants whose duty it is to monitor, at times, they feel it is an added duty for them”* [RA, FGD 1]

Other personal attributes [characteristics of individuals] of family members facilitated use of the intervention in unexpected ways. Neighbouring patients and their families would assist others who were facing difficulty. Family members took pride in their ability to help with the responsibility of monitoring. This encouraged them to do what they were asked.

*“…*..*they knew each other, so she was very proud, she said ‘you hurry, I take your observations, hurry hurry position yourself*.*’ She was saying ‘now I am going home when I think I am now also a nurse’ she was very proud”* [Nurse, FGD 2]

### Barriers

Self-efficacy [characteristics of individuals] describes an individual’s belief in their own capabilities to perform the necessary actions to achieve the tasks set out in the intervention. Family members supporting their patients in hospital come from a large range of backgrounds. Some are educated with professional jobs and others are illiterate and unable to read and write. Almost half of the 315 participants excluded from the trial were due to illiteracy. The impact of this on the ability of family members to be involved was described clearly during the focus group discussions. However, despite this, other family members demonstrated willingness to learn and be involved because they recognised the importance of monitoring their patients closely.

*“I got one and she couldn’t write, but she could interpret so was taught that if the oxygen levels are ok put a tick, if they are not ok put a cross. She was able to count the breathing but writing the real figure was a challenge*.*”* [RA, FGD 1]

A second identified barrier was the challenge presented by the available resources [inner setting]. In a low-income setting there are usually less staff available and less access to equipment and supplies within the hospital. Patients were deemed sick enough to require continuous monitoring, preventing use of the pulse oximeter by family members for routine monitoring of their postoperative patients.

*Nursing staff were approached and asked to take the machine in the general ward but they refused giving a reason that they have a right to every equipment and the pulse oximeter should be on that patient for close observation* [Field notes]

After identifying abnormal vital signs, family members were trained to notify a member of staff so that they can intervene however, limited human resources meant that at times there were no members of staff available on the wards. An example would be a single nurse on duty who may be required to carry out duties off the ward, for example collecting supplies from stores. Nursing staff available to respond appropriately to communication from family members was an assumption that we made in our programme theory. Necessary absence from the ward was one example when this assumption failed. A second example were the cultural attitudes [inner setting] identified in some members of staff, patients and their family members. With low salary scales and poor remuneration for work, it is common for staff to work more than one job. Staff on duty at night routinely sleep and family members were reluctant to wake them. This potentially creates a barrier preventing family members from seeking help when they need to.

Other cultural norms within the context of the hospital where the pilot trial ran were also noted to create barriers to use of the intervention. Research in Uganda is commonly associated with incentives for participants who are recruited. This has created a culture of expectation.

*Family members who do the vital signs, think the organisation should give them ‘something’, probably cash or incentives for the work they are doing*. [Field notes]

Complexity [intervention characteristics] of an intervention plays a large role in its ease of implementation. Although the respiratory rate is the best early predictor of patient deterioration, globally it is also the vital sign that is most frequently missed out.^17^ Measuring a respiratory rate accurately requires several steps. Recognition of each individual breath a patient is taking, counting the number of breaths and simultaneously watching the clock. Family members found difficulty coordinating both timekeeping and counting at the same time and for those that chose to use their phones instead of the clock to keep time, simple ‘button’ phones presented a challenge.

Other aspects of the routine workflow in the regional hospital where the SMARTER pilot trial was conducted commonly led to challenges in the intervention implementation. We coded these barriers under compatibility [inner setting]. Examples include exclusion of family members from the entire clinical areas to allow medical student examinations to take place, preventing them from taking vital signs, and patients unexpectedly going home directly from theatre despite being recruited, family members trained and being expected to return to the ward after surgery. In the obstetric department, particularly on the postnatal wards the high throughput of patients led to continuous movement around the ward disrupting the recording of vital signs. There were also common changeovers of family members. Patients who undergo their operations at night often have different caretakers the following day. This was particularly true for the obstetric patients.

*Some caretakers bring in their mothers or sisters to give birth, they are learned and willing to enrol in the study, so they consent. However, in the morning, the consented caretaker disappears and leaves [an] elderly grandmother or mother-in-law to take care of the patient. The grandma or mother-in-law can’t write her name and…*..*only speaks a local language*. [Field notes]

### Pre-defined intervention evaluation

Feedback gained during our pre-planned review identified areas of complexity that were able to be resolved through amendments and resubmission and approval by the local research and ethics committee. These are summarised in table 2. Differences in the order of documenting vital signs on the chart compared to the numbers visible on the pulse oximeter led to incorrect documentation of vital signs. (Figure 2) The use of symbols instead of words on the vital signs chart was found to be confusing and in the local language, the times of day were described differently. Version 1 of the observation chart listed the times of day using the 12-hour clock with the day beginning at 12am.

**Table 2.** Summary of changes made to the intervention following the pre-specified evaluation at 2 months.

| **Intervention area** | **Original intervention** | **Amended intervention** | **Reason for change** |
| --- | --- | --- | --- |
| Training of family members | Training planned twice a day in groups | Training conducted when family members available, individually or in groups | Family members never available on the ward at the same time, scheduling for surgery at chaotic at times |
| Family member documentation of vital signs | Time of day listed as 12-hour clock using numbers only | Times of day as understood in local culture included (eg. breakfast) | In the local culture, the day begins at 6am and so 8am ('8 hours after the beginning of the day') would equate to 2pm. Local language incorporated to avoid confusion |
| Family member documentation of vital signs | Use of > and < signs to mean greater or less than | Words used instead of symbols | Family members and patients did not understand the symbols |
| Family member documentation of vital signs | Choice of four ranges when documenting respiratory rate and heart rate | Simplified to three ranges, above the upper limit, below the lower limit and normal | No added benefit to two options within the specified 'normal range', additional options led to some confusion |
| Family member documentation of vital signs | Heart rate above respiratory rate in the observation chart | Order changed round to match the numbers visible on the pulse oximeter with respiratory rate above the heart rate | This simplification enabled more accurate differentiation between heart rate and respiratory rate |

**Figure 2.**
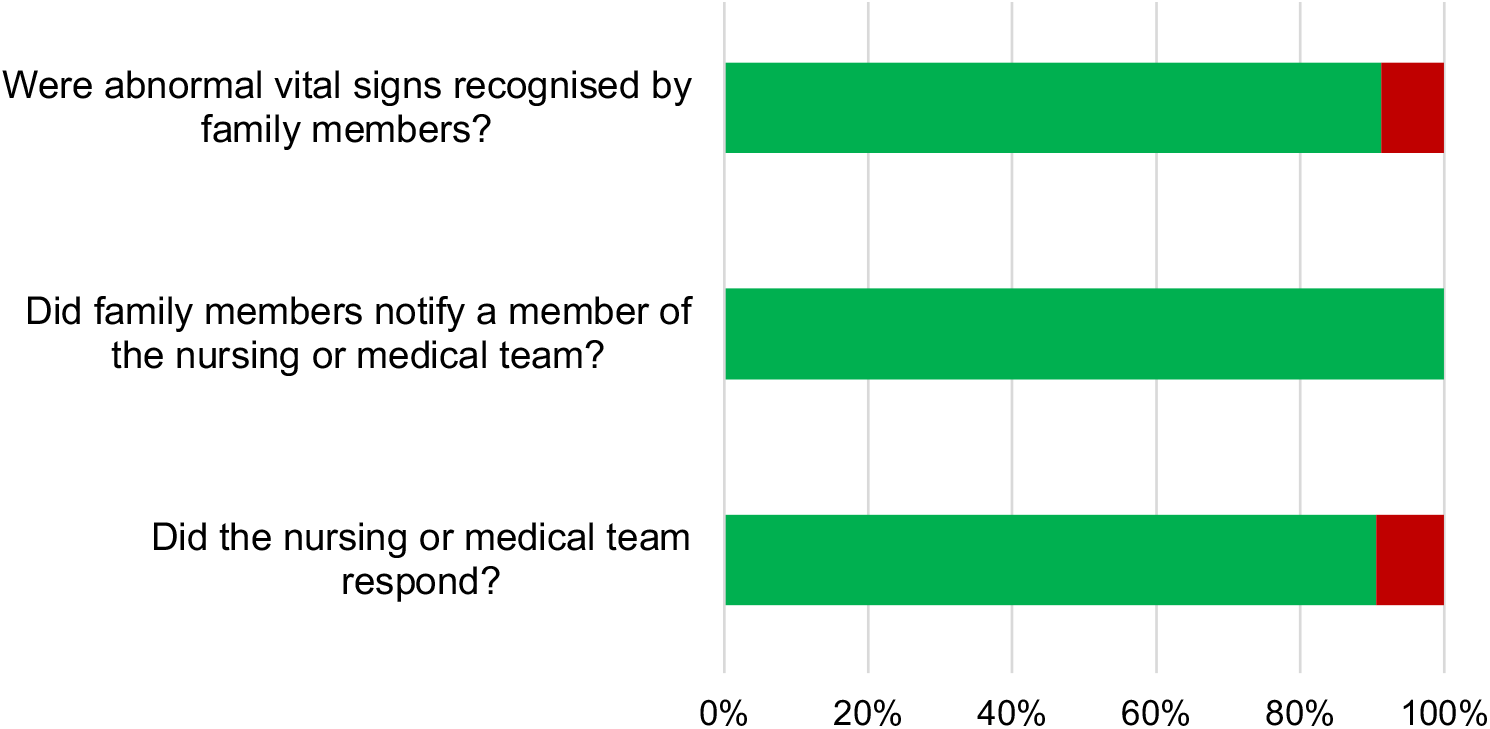
Family member and staff response to abnormal vital signs during the SMARTER intervention. Green = yes.

**Figure 3.**
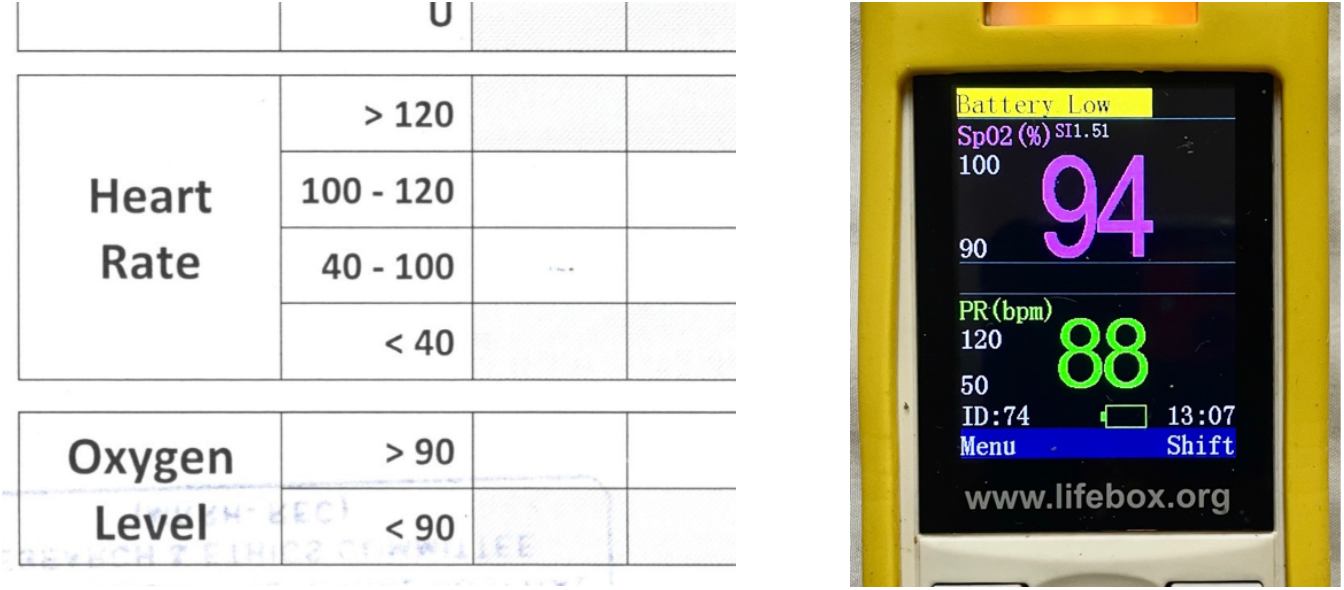
Images of the first version of the family member observation chart and the pulse oximeter used, illustrating the discrepancy between positioning of heart rate (bottom, in green on the pulse oximeter) and oxygen level (top, in pink on the pulse oximeter)

Locally however, the day begins at 6am. 11am in the 12-hour clock would be interpreted as 5pm in the local language. To overcome this, the times of day were described using common activities e.g. breakfast or lunch and the local translations of these times were used. Observation charts used in the SMARTER pilot trial can be found in the supplementary material of the main trial publication.

## Discussion

Our process evaluation provides important contextual understanding to support the SMARTER pilot trial results. In a subset of patients, most sets of abnormal vital signs were recognised by family members, all of those recognised were communicated to a member of nursing or medical staff and the staff responded most of the time. Themes that facilitated the family supplemented vital signs monitoring intervention included the relative advantage, relative priority and tension for change. Individual characteristics centred around knowledge and beliefs also resulted in a positive influence. Major constructs from the consolidated framework for implementation research that had a negative influence included available resources, local culture and compatibility of the intervention.

In 2020, Leonard et al. published a systematic review of barriers and facilitators to implementing health innovations in LMICs.^18^ Although a different conceptual framework was used to assimilate the results from the systematic review, it is easy to identify cross-cutting themes that match those identified in our process evaluation and that contribute to a facility’s implementation readiness. Examples include resource limitations (for example human, physical and time) and contextual factors such as socio-cultural influences.^18-21^ van den Hoed et al. use a scoping review to describe four main factors and 10 sub-factors that contribute to an organisation’s innovation readiness.^22^ Innovation readiness in this context can be considered synonymous with implementation readiness. Whilst these small differences in nomenclature exist, the overarching theme remains. A lack of facility or institution readiness for innovation will act as a barrier to implementation success. Understanding influences that affect the introduction of a clinical intervention is crucial to help inform future clinical trials and work to scale implementation at a regional or national level.^23^ At Mbale RRH, there is a tension between the implementation climate, illustrated by the healthcare worker’s recognition of gaps in postoperative monitoring and their desire to change; and the ability of the teams on the ground to put this into practice (implementation readiness). Damschroder et al. describe six sub-constructs that contribute to a positive implementation climate for an intervention.^16^ The first, tension for change, describes dissatisfaction that staff have with current processes. This tension is difficult to create if it does not already exist.^24^ At Mbale RRH where the nurse to patient ratio can be as high as 1:40,^2^ the staff recognise that current levels of postoperative monitoring are not adequate and this drives their desire for something different to happen to address this gap. Alongside tension for change, successful implementation is supported by a shared belief that the intervention is important (relative priority), that it works, and that implementation will result in an advantage for the healthcare workers involved (relative advantage). These allow staff to prioritise the intended intervention when their workload is overwhelming. The relative advantage of innovations to improve monitoring of patients has also been described in several other countries.^25-27^ In West Africa, clinicians recognised the added advantage of wearable sensor technology in paediatric patients, especially on wards where current levels of monitoring equipment were limited.^27^ In HICs, improved postoperative monitoring using novel technology has shown benefit both in the community and before hospital discharge.^28,29^ Implementing technology driven innovations in low resource settings in a sustainable way is however, much harder, facing significant challenges within the enabling environment, existing infrastructure and integration with existing systems.^30^ Whilst the SMARTER Pilot Trial did not focus on the use of technology to improve postoperative monitoring, the barriers that affect implementation readiness for a new innovation cut across health systems.

The complexity of a healthcare system necessitates a formal process evaluation to run alongside complex intervention research. Our pilot trial is strengthened by its co-design with a formal process evaluation. We followed established guidance for the evaluation of complex interventions providing robust context to support future efforts to test the SMARTER intervention on a larger scale. We highlight important themes that influence implementation work in low-resource environments that can be used by others during the design phases of their implementation development. Weaknesses of our process evaluation include limited stakeholder involvement in the design of the process evaluation itself. Future work should involve patients and their carers from the time of study conception. Patient researcher led focus group discussions with patients and their family members would be one way to address this gap in the future. The process evaluation was based on a relatively small pool of qualitative data. Although field notes were kept by the research team throughout the six-month duration of the trial, only two focus group discussions were held. The process evaluation and main pilot trial occurred during the COVID-19 pandemic and gatherings of groups of people was limited. Whilst we believe we reached thematic saturation by combining data from the field notes and focus group discussions, there is a risk that this may not be the case. We did not set out to test intervention effectiveness from the study start, this was added after our predetermined evaluation at two months and after protocol amendments were approved. These delays resulted in a relatively small sample of quantitative data.

## Conclusions

Process evaluation of the SMARTER pilot trial provides valuable insight into the context within which it was tested. Family members were able to identify abnormal vital signs and communicate them to the healthcare team. Prespecified review allowed us to make improvements to the intervention. Barriers and facilitators influencing trial success in this context have been described and this learning must be incorporated in the design of future, larger trials to test the effect of the intervention on clinical outcomes.

## Data Availability

All data produced in the present study are available upon reasonable request to the authors

## Data sharing

Requests for data sharing are welcomed from bona fide researchers.

## Acknowledgements

The authors would like to extend their thanks to all the patients, family members and staff at Mbale Regional Referral Hospital who were involved in this pilot study.

## Authors’ Contributions

AHS: Study conception and design, data analysis, writing first draft of paper; FB: Study conception and design; JuN, JoN, JK, HAM, LN, MK, WW, CM, MS, BE: Patient recruitment, data collection; TS: Study conception and design, data analysis; RP: Study conception and design. All authors reviewed, revised, and approved the final article.

